# Disability and Post-traumatic Stress Symptoms in the Ukrainian General Population During the 2022 Russian Invasion

**DOI:** 10.1101/2022.11.07.22282027

**Authors:** Tarandeep S. Kang, Robin Goodwin, Yaira Hamama-Raz, Elazar Leshem, Menachem Ben-Ezra

## Abstract

**Background:** Previous research has shown that people with disabilities are disproportionately vulnerable to symptoms of psychological distress after exposure to armed conflict. Past work has also shown that individuals displaced by conflict are at heightened risk of post-traumatic stress. However, we know of no research that has attempted to examine the association between disability severity and post-traumatic stress symptoms in a general population affected by armed conflict.

**Method:** We examined the association between levels of functional disability in the Ukrainian population and symptoms of post-traumatic stress during the 2022 Russian invasion of Ukraine. We analysed data from a national sample of 2000 participants from across this country, assessing disability using the WHODAS-12 (six domains of disability) and the ITQ assessment of ICD-11 PTSD symptomatology. Moderated regression examined the impact of displacement status on the disability-post-traumatic stress relationship.

**Results:** Different domains of disability predicted post-traumatic stress symptoms to varying extents, with overall disability score significantly associated with post-traumatic stress symptoms. This relationship was not moderated by displacement status. Consistent with previous research, females reported higher levels post-traumatic stress.

**Conclusions:** In a study of a general population during a time of armed conflict individuals with more severe disabilities were at greater risk of post-traumatic stress symptoms.

Psychiatrists and related professionals should consider pre-existing disability as a risk factor for conflict -related post-traumatic stress.

**Highlights:** 

**What is already known on this topic:** People with disabilities are vulnerable to a variety of forms of psychological distress after experiencing disasters of various kinds, including armed conflict. However, associations between levels of disability in the general population and post-traumatic stress during a period of conflict have not previously been examined.

**What this study adds:** Increasing severity of functional disability in the general population is associated with increasing post-traumatic stress symptoms in a conflict exposed population.

**How this study might affect research, practice or policy:** Mental health professionals dealing with conflict affected populations should take account of the additional vulnerabilities caused by disability status, and work to mitigate this by providing access to functional and social support.

## Introduction

Conflict between Russia and Ukraine has been ongoing since 2014, with initial hostilities in Crimea and Donbas followed by a wider Russian invasion in February 2022. As has been seen in other military conflicts, this has had a serious deleterious impact on the mental health of affected populations [1]. The ongoing conflict has also led to the substantial displacement of populations within Ukraine, with this population at risk of additional burden on their psychological health. A national survey of Ukrainian internally displaced people (2016) reported a PTSD prevalence of 32%, 22% for depression 22%, and 17% for anxiety [2].

People with disabilities can face particularly significant negative mental health consequences after disaster and are at considerable risk following war related trauma. A study of elderly individuals conducted in 2016, during a period of active conflict in the aftermath of the annexation of Crimea [3], found 33.6% of respondents living in Ukrainian government-controlled areas, and 42.5% of respondents living in regions not under Ukrainian control showed symptoms of severe psychological distress using the default threshold for the Kessler K6 scale [4]. Women, those who were more dependent on others for care and support, and those with chronic health conditions and disabilities were at the greatest risk of psychological distress. Data collected in 2002 from the Afghan population [5] found that 71.7% of respondents with disabilities reported depression, 84.6% anxiety (both using the Hopkins Symptom Checklist-25 [6,7]) and 42.2% PTSD (using the Harvard trauma questionnaire [8]). Overall, women reported worse mental health than men. In a subsequent study of Afghan respondents (2004-2005) [9] 86.1% of those with disabilities showed symptoms of potential mental distress using default K6 criteria, with greater prevalence of distress amongst women.

Despite this evidence of psychological trauma amongst the Ukrainian population, and the high risk to mental health of people with disabilities exposed to armed conflict, we know of no previous work that has examined the association between disability severity and post-traumatic stress symptoms in a general population affected by armed conflict. To this end, we assessed associations between disability and posttraumatic stress symptoms in the population employing a national sample from the Ukrainian population. We collected data in the first weeks after the invasion, during which attacks from Russia occurred across the whole country. We hypothesise that (a) severity of disability will predict severity of post-traumatic stress symptoms and (b) females will experience higher rates of post-traumatic stress symptoms [10]. Previous studies indicate that internally displaced people and refugees are systematically more vulnerable to various forms of psychological distress, than people who have not been displaced in conflict [11,12] and we expect to replicate this trend. We also explore the extent to which this displacement will moderate any association between disability and post-traumatic stress.

### Methodology

#### Recruitment and Sample

We report data from a national sample of 2000 people aged 18-55 surveyed five weeks after the start of the invasion (M age 37.18 years, SD= 9.3), 1026 (51.3%) females [14; for further details]. Participants were drawn from a panel maintained by the survey company Kantar, Ukraine to provide general population representativeness for age, gender, and pre-displacement region of the country. Sample descriptives are presented in Table 1.

**Table 1:** Descriptive statistics

| Variable | M (SD) | Frequency (%) |
| --- | --- | --- |
| Age | 37.18 (9.23) |  |
| Sex (Male) |  | 974 (48.7) |
| Sex (Female) |  | 1026 (51.31) |
| Displacement |  | 1455 (72.8) |
| Displaced |  |  |
| Not Displaced) |  | 545 (27.2) |
| ITQ Score (Post-Traumatic Stress Symptoms) | 11.64 (4.91) |  |
| WHODAS Domain (range 1-5): Getting Around | 1.71 (1.78) |  |
| WHODAS Domain: Self-Care | 1.07 (1.67) |  |
| WHODAS Domain: Getting Along with People | 1.71 (1.81) |  |
| WHODAS Domain: Life Activities | 2.00 (1.82) |  |
| WHODAS Domain: Participation in Society | 2.17 (1.87) |  |
| WHODAS Domain: Understanding and Communicating | 1.83 (1.77) |  |
| WHODAS Total | 10.51 (8.65) |  |

All participants provided informed consent. Ethical approval for original data collection was provided by the Ethics Committee of the final author; approval for this re-analysis by the Ethics Committee of the first author. All aspects of this research were performed in accordance with the principles of the Declaration of Helsinki [14]. The dataset is available on the Open Science Framework (https://osf.io/z5adg/).

#### Measures

*Post-traumatic stress symptoms (PTSS)* were assessed using six items from the International Trauma Questionnaire (ITQ) [15] previously validated for use with the Ukrainian population [16]. Participants were asked how much they were bothered by each of six symptoms (e.g. “feeling jumpy or easily startled”) in the previous month and responded on a five-point Likert scale ranging from not at all (0) to extremely (4) (α 0.86). Possible scores ranged between 0 - 24.

*Disability status* was measured using the 12 item WHODAS 2.0 [17] which has previously demonstrated reliability and validity in international use [18] although only the full 36-item version has been previously used in the Ukrainian context. The 12 items measure difficulties in six different domains of functioning, with two items per domain: 1: Cognition; 2: Mobility; 3: Self-care; 4: Getting along with others; 5: Life activities; 6: Participation (overall α = 0.91). Respondents indicated the level of difficulty they had with each item using a five-point Likert scale, from none (1) to extreme or cannot do (5). We used the simple scoring method [17] adding together scores to create a single overall score (from 12 (no disability) to 60 (complete disability).

To assess displacement respondents were coded as *displaced* if either they had been forced to move within Ukraine as result of the conflict or had to leave the country because of the war.

#### Analyses

To assess associations between disability and PTSS we initially ran Pearson correlations between each subscale of the WHODAS-12 and the total post-traumatic stress score from the ITQ. We then ran a moderated regression with WHODAS-12 total score as the focal predictor and post-traumatic stress score as the outcome. Displacement status was used as the moderator, with sex as a covariate. All analyses were conducted using SPSS version 27. Moderated regression was conducted using the process macro [19] with a heteroscedasticity-consistent standard error estimator [20].

## Results

Mean post-traumatic stress symptoms score was 11.64 (range 0-24, SD= 4.91), total disability (WHODAS-12) score was 10.51 (range 12-60, SD = 8.65). Average WHODAS-12 (disability) subscale scores ranged from 1.07 (SD = 1.67) for self-care to 2.17 (SD = 1.87) for participation in society. There were moderate [21] positive zero-order correlations between each of disability measure of post-traumatic stress symptoms All six of the disability sub-scales were significantly associated with PTSD symptoms (rs 0.30 (self-care) -0.45 (participation in society), all p< .001) (Table 2).

**Table 2:** Correlations Between WHODAS Subscales And ITQ Total Score

|  |  | WHODAS<br>Getting<br>Around | WHODAS<br>Self-Care | WHODAS<br>Getting Along<br>With Others | WHODAS<br>Life Activities | WHODAS<br>Participation In<br>Society | WHODAS<br>Understanding &<br>Communicating |
| --- | --- | --- | --- | --- | --- | --- | --- |
| WHODAS Self-Care | r | .56** |  |  |  |  |  |
|  | Sig. | .000 |  |  |  |  |  |
|  | N | 2000 |  |  |  |  |  |
| WHODAS Getting<br>Along With Others | r | .45** | .56** |  |  |  |  |
|  | Sig. | .000 | .000 |  |  |  |  |
|  | N | 2000 | 2000 |  |  |  |  |
| WHODAS Life<br>Activities | r | .60** | .58** | .59** |  |  |  |
|  | Sig. | .000 | .000 | .000 |  |  |  |
|  | N | 2000 | 2000 | 2000 |  |  |  |
| WHODAS Participation<br>In Society | r | .58** | .52** | .57** | .62** |  |  |
|  | Sig. | .000 | .000 | .000 | .000 |  |  |
|  | N | 2000 | 2000 | 2000 | 2000 |  |  |
| WHODAS<br>Understanding &<br>Communicating | r | .59** | .56** | .56** | .68** | .68** |  |
|  | Sig. | .000 | .000 | .000 | .000 | .000 |  |
|  | N | 2000 | 2000 | 2000 | 2000 | 2000 |  |
| ITQ Total | r | .34** | .30** | .34** | .40** | .45** | .43** |
|  | Sig. | .000 | .000 | .000 | .000 | .000 | .000 |
|  | N | 2000 | 2000 | 2000 | 2000 | 2000 | 2000 |

\*\* . Pearson r. Correlation is significant at the 0.01 level (2-tailed).

In line with our first hypothesis, a moderated regression model (Table 3) indicated that increasing levels of functional disability in the Ukrainian population predicted greater endorsement of post-traumatic stress symptoms (β = 0.27, t = 21.67, p = < .001). Also, as anticipated, displacement was significantly associated with post-traumatic stress symptoms although the disability-post-traumatic stress symptoms relationship was not moderated by displacement (β = -0.01, t = -0.43, p = .67). Women exhibited more post-traumatic stress symptoms (β = 1.71, t = 8.88, p = < .001).

**Table 3:** Moderated linear regression on disability and post-traumatic stress symptoms

| Predictor | $\beta$ | Standard error | t | *P* | 95% confidence interval |
| --- | --- | --- | --- | --- | --- |
| Constant | 8.83 | .54 | 16.40 | <.001 | 7.78, 9.89 |
| Sex | 1.71 | .19 | 8.88 | <.001 | 1.33, 2.08 |
| Disability | .27 | .01 | 21.67 | <.001 | .22, .27 |
| Displacement | .43 | .15 | 2.83 | .0046 | .13, .73 |
| Interaction (disability x displacement) | -.01 | .02 | -.43 | .66 | -.05, .03 |

## Discussion

Ukraine was faced with substantial mental health challenges even before the Russian invasion in 2022. Data collected in 2002 for the Ukraine World Mental Health Survey found lifetime mental illness prevalence of 31.6% and one of the highest suicide rates in Europe [22].

Previous research has shown that people with disabilities are particularly vulnerable during times of conflict [5,9]. In this paper we present one of the first analyses to examine associations between disability severity and post-traumatic stress symptoms during a period of war. We find that that women and people with more severe disabilities endorsed greater post-traumatic stress symptoms, as do those who are displaced, but there was no moderation of the disability-post-traumatic stress relationship by displacement.

A variety of disabilities including vision impairment [23] physical disability [24] and autism spectrum disorders [25] have each been previously associated with increased vulnerability to PTSD. The associations between post-traumatic stress symptoms and all six dimensions of the WHODAS 12 suggest that increasing disability severity may make individuals more vulnerable to post-traumatic stress during a period of conflict. At the same time, there were differences in associations between disability and post-traumatic stress symptoms by disability domain. Mobility difficulties were the most weakly associated with post-traumatic stress, while impairments related to social functioning had the strongest association. This may be because respondents with more severe physical/intellectual impairments may not be available when sampling the general population, given that the Ukrainian health and social care system is most likely to support these people in an institutional setting.

Previous research [12,26] has suggested that refugees and internally displaced people are at heightened risk for a variety of mental health problems when compared to individuals who have not been displaced. While we did find that displacement status was associated with post-traumatic stress symptoms, it did not have a moderating role in the disability-post-traumatic stress relationship. This may be because of a lack of possibility for movement amongst refugees with disability. While housing, employment and access to health and support services were already problematic for Ukrainians with disabilities living in the Donetsk and Luhansk Oblasts in the east of the country [27] it may have been particularly difficult for people with disabilities in these populations to move. Indeed, previous research on predictors of mobility among Ukrainians after the annexation of Crimea in 2014 [28] finds that people with disabilities were less likely to leave their homes than their nondisabled counterparts. Our finding that female gender is associated with greater endorsement of post-traumatic stress symptoms is consistent with previous studies of populations exposed to war related trauma [29].

Our study has several strengths. We present a novel examination of disability and PTSD during conflict employing the widely used the WHODAS 2.0 12-item measure of disability. We note that our disability scores measured using WHODAS 2.0 in the Ukrainian population five weeks after the Russian invasion were only slightly higher than previously established norms for the Polish population using the same scoring method, during peacetime [30]. This suggests that our measure accurately depicts the prevalence of pre-existing functional impairment in the Ukrainian population and is not overly biased by the presence of newly acquired disabilities caused by the conflict.

At the same time, we recognise several limitations. Firstly, this study was of the Ukrainian general population. There are substantial differences in the treatment of people with and without disabilities in Ukraine. As a result of the Soviet past – and continued influence of Soviet and Russian traditions even following the independence of Ukraine – people with disabilities do not have equal rights [31] with many placed in long-term institutional care [32]. This means that people with especially severe disabilities who we might infer would be at the highest levels of risk from PTSD after being exposed to the war were largely inaccessible for study. Second, because we assessed PTSS in the early weeks of the Russian invasion, we focused on post-traumatic stress symptoms using the ITQ (ICD-11), but did not assess PTSD symptoms prior to the invasion. Third, we did not explicitly ask about types of disabilities, meaning we were unable to identify any specific disabilities associated with PTSD symptomology. Finally, we excluded participants aged over the age of 55 due to their more limited use of the internet in Ukraine. This nevertheless restricted our analyses of age effects on PTSD symptoms. Finally, we did not measure PTSD symptoms prior to the invasion.

Our study has several implications for practitioners. First, we add to the existing literature on war trauma and disability by showing that a range of disability/functional impairments may place patients at risk of post-traumatic stress. Given this range of risk factors health professionals should take account of the broader infrastructure and environmental damage caused by conflict and its implications for social functioning as well as daily care for many people with disabilities [33]. Practitioners need to make use of evidence-based strategies for improving psychosocial health in emergencies [34], with particular reference to the unique needs of people with different disabilities. In this case medical personnel may also wish to direct people with disabilities to services and support groups that not only assist in meeting their basic needs but provide opportunities for building or augmenting a social support network. This support is likely to be important for people with disabilities in general [35] and those relocated as a result of conflict [36,37].

## Data Availability

All data produced used in this study are available online at the OSF.

https://osf.io/z5adg/

## Data Availability

Scientists can obtain access to individual-level data from the UK Biobank by applying to UK Biobank (https://www.ukbiobank.ac.uk/enable-your-research).

